# Trends in fluoroquinolone prescribing in UK primary and secondary care between 2019 and 2023

**DOI:** 10.1101/2024.03.04.24303729

**Authors:** Fergus Hamilton, Elizabeth Darley, Alasdair MacGowan

## Abstract

**Introduction:** Fluoroquinolones are important, widely used antibiotics but have associations with a significant number of adverse outcomes. A recent (January 2024) decision by the UK drug regulator, the Medicines and Health Regulatory Authority (MHRA), restricted systemic use of these antibiotics to when only absolutely necessary with immediate effect. One stated reason for the ban was the failure of previous guidance (2019, 2023), to impact prescribing, with the 2023 MHRA Drug Safety Update stating there had been “no change in prescribing” of fluoroquinolones in relation to guidance.

**Methods:** We evaluated the trend in prescribing of fluoroquinolones and comparator antibiotics using national data for all primary care practices in England from 2019 to 2023. We calculated the percent change in prescribing of fluoroquinolones using linear regression, comparing with other antibacterials Analyses were then performed at the integrated care board (ICB) level. We also performed a similar analysis on secondary care prescribing and included hospital inpatient stay data.

**Results:** In primary care, there was a clear negative trend in fluoroquinolone (particularly ciprofloxacin) item dispensing, with a 4.2% reduction in items dispended per year, 95% confidence interval, CI (−5.2%; −3.3%, p = 6 x 10^-13^). This occurred despite no change in overall antibacterial prescription (+2% −0.56%; +4.6%, p = 0.12) and no decrease in comparator antibiotics. These occurred across nearly all (97/101) ICBs. Secondary care data showed stable prescription of fluoroquinolones, but other comparator antibiotics increased, leading to relatively fewer prescriptions compared to other agents.

**Conclusions:** There was a marked reduction in fluoroquinolone prescribing in primary care in England in both absolute terms and relative to other antibiotics between 2019 to 2023. Relative reductions have occurred in secondary care.

**Funding:** Wellcome Trust (222894/Z/21/Z)

## Introduction

Fluoroquinolones are a widely used class of antibiotics.^1^ They are effective therapy for Gram-negative, Gram-positive, and (for some agents) anaerobic bacteria.^1^ The most commonly used agents are ciprofloxacin, levofloxacin, and moxifloxacin, although the first two are far more common. Given their broad spectrum of in-vitro activity and generally favourable pharmacokinetics, they are widely recommended across a broad range of infection indications, particularly in patients with penicillin allergy. They were (until very recent MHRA announcements^2^) the recommended option for severe community acquired pneumonia (CAP) with a penicillin allergy in UK National Institute for Clinical Excellence (NICE) guidance^3^. They are also recommended in the 2019 Infectious Diseases Society of America/American Thoracic Society (IDSA/ATS) pneumonia guidance,^4^ and as first line and/or recommended in penicillin allergy or under other conditions in conditions as broad as prosthetic joint infection, legionella pneumonia^4^, sexually transmitted infections^5^, typhoid fever^6^, pyelonephritis^7^, tuberculosis^8^ and Gram-negative infections including bacteraemia^9^. For some deep tissue infections they may be given for a course lasting several weeks or months, and are the preferred therapy. Given the paucity of effective oral agents for Gram negative infection and high resistance rates to other drugs, fluoroquinolones are highly valuable agents. For infections caused by some organisms such as *Pseudomonas aeruginosa*, they are the only oral option.

Despite these benefits, fluoroquinolones are known to associate with a range of adverse events.^10^ Well established and proven causal associations include tendonitis and *C. di0icile* infection, but over the past 20 years, an increasing number of associations have been identified including QTc prolongation, aortic dissection, aortic aneurysm and (sometimes severe) psychiatric associations.^10^ Regulators have been aware of these risks for some time, with the first FDA ‘black box’ warning in 2008 (related to tendonitis).^11^ More recently the European Medicines Agency (EMA) and MHRA have produced multiple updates with increasing concern. The EMA produced guidance suggesting use should be limited to only severe infection in March 2019 (due to the risk of tendonitis and potentially aortic dissection).^12^ The MHRA have also produced a number of updates, starting in 2019^13^, repeated in 2023^14^, and most recently, in 2024^2^, a much tighter restriction of fluoroquinolone prescribing. This directs against any use of systemic fluoroquinolone therapy except where absolutely necessary (e.g. no other drug can be used). The extent of this ban resulted from an ongoing review of the evidence, increasing patient reports via the national scheme (Yellow Card), and “no evidence of a change in fluoroquinolone prescribing patterns” in relation to previous warnings^14^.

The reference for this “lack of change” in prescribing was a European pharmacovigilance study^15^, which included 15 million UK participants from 2016 to 2020 and was commissioned by the EMA to assess the impact of the 2019 EMA guidance across numerous countries. It was not able to identify a reduction in fluoroquinolone usage directly in relation to the EMA guidance.^15^ However, this study did show that a) the UK had the lowest rates of fluoroquinolone prescribing in all included countries, and b) the UK had around a 3% reduction per year in fluoroquinolone prescriptions from 2016 to 2020 guidance.^15^

Given this study ended during 2020 (when impact of the COVID-19 strongly impacted antimicrobial prescribing) and showed an apparent reduction in prescribing we performed a study of UK fluoroquinolone prescribing in primary care using all of England primary care and secondary care data. Our primary aim was to identify the recent trend in fluoroquinolone prescribing.

## Methods

To evaluate trends in prescribing of fluoroquinolones, we extracted data on England wide prescribing in both primary and secondary care datasets.

### Primary care dataset

Primary care data was accessed via OpenPrescribing^16^, which extracts directly from NHS primary care records for England. We extracted specific data on quinolones (as a class), ciprofloxacin, and levofloxacin. Fluoroquinolones are the only regularly prescribed quinolones in the U.K and so we use the terms interchangeably in this report. Ciprofloxacin accounted for >95% of prescriptions, and no other quinolone agents were prescribed widely.

For comparison, we also extracted data on two comparator agents with similar spectrums of activity: co-amoxiclav and co-trimoxazole. We also extracted total antibiotic use for all penicillins, all antibacterials, and total medication use. Data was available from December 2018 until November 2023 (4 years 11 months), at a monthly level, and at Integrated Care Board level. There are 101 Integrated Care Boards in ?England. Data is in the format of number of prescriptions and quantity dispensed (as most prescriptions are for more than one dose). We estimated defined daily doses (DDDs) using BNF dosing (i.e. as ciprofloxacin is dosed twice daily), the quantity divided by two is the DDD.

### Secondary care dataset

Secondary care data was accessed via (https://hospitalmedicines.genomium.org/)^17^ which in turn extracts data from the NHSBSA SCMD dataset for England.^18^ We extracted data on ciprofloxacin and levofloxacin, as well as comparator agents with similar spectrum of activity: co-amoxiclav, piperacillin-tazobactam, co-trimoxazole, meropenem, clarithromycin, amoxicillin, and ceftriaxone. As this dataset reports chemical products, we defined piperacillin-tazobactam by tazobactam sodium, co-trimoxazole by sulfamethoxazole, and amoxicillin-clavulanate by potassium clavulanate. None of these are widely used outside the antibiotic combination. Data was available from April 2019 until November 2023, at a monthly level.

We estimated the number of prescriptions by assuming a 5-day course length for all infections and assuming BNF standard dosing regimens. This led to one prescription being 5g for Ciprofloxacin, 5g for levofloxacin, 15g for Meropenem, 15g for Amoxicillin, 5g for clarithromycin, 5g for Ceftriaxone 8g for co-trimoxazole (assuming 800mg per dose of sulfamethoxazole), 3g for co-amoxiclav (assuming 200mg per dose of clavulanate, and IV therapy), and 7.5g for piperacillin-tazobactam (assuming 500mg per dose of tazobactam). We recognise these are assumptions and are simply produced to aid comparison with primary care prescribing.

### Hospital data

From December 2020 until December 2023, NHS England produced monthly hospital utilisation data. This is in the format of number of occupied bed days per month and forms part of the Urgent and Emergency Care Situation Report.^19^

### Analytic approach

As there have been numerous warnings from multiple drug regulators including the FDA, MHRA, and EMA over this time, and as the interventions are likely not applied instantaneously, we took the approach of visualising and quantifying change over time, rather than performing an analysis at a specific time point. This approach has also been shown to be highly sensitive to statistical methods and/or time points chosen.^20^ Our datasets largely overlap although the first major MHRA warnings and restriction occurred in March 2019, prior to our secondary care dataset.

For this reason, and because of the increased potential for potentially less appropriate prescribing occurs in primary care (due to the reduced severity of illness and reduced burden of bacterial infection, and less use of fluoroquinolones in guidelines), we focus most of our analysis on the primary care data but include the secondary care data for completeness.

We performed linear regression for each drug/drug class with date as the exploratory variable, with the scale set to evaluate changes in prescriptions per year. Absolute prescription rates were calculated with respect to the total English population in the given year. To aid visualisation, we scaled some analyses so prescriptions on the first study month were the reference, and report figures as a percentage change. As regional data (from ICBs) was available for primary care data, we also ran analyses at the ICB level.

To compare with other classes/antibiotics, we performed a Z-test for the beta and standard error of the linear regression for each class.^21^ For aid of visualisation, we also performed linear regression of ratios (e.g. the ratio of co-amoxiclav to ciprofloxacin over calendar time).

Finally, for the time where it was available, we analysed data adjusted for inpatient population. That is, we divided the number of prescriptions by the number of inpatients for each month to generate prescriptions/month/patient.

### Software and codes

We used R v 4.31 for analyses, and used the *tidyverse* package for data wrangling and plotting.^22^ All code is available at https://github.com/gushamilton/floroquinolone, and the analysis is completely replicable by running the same code locally. Regression was performed using the linear regression model in R, while some plots fitted splines via ggplot.

### Ethics

All data used in this analysis is publicly available. No ethics approval was therefore required.

### Funding

FH’s time was funded by the Wellcome Trust.

## Results

Our primary analysis focussed on primary care prescribing, where the risk-benefit ratio of fluoroquinolone prescribing is most questioned. Across the study period (∼5 years), 151,144,401 prescriptions for antibacterial agents were given across England, with an estimated population of 59.6 million (2021 census data^23^). This equates to 549 prescriptions per year/1000 people. Approximately one fifth of this was made up of amoxicillin (125 prescriptions per year per 1000 population). Fluoroquinolone represented a small proportion of antibiotics prescribed (9 per year per 1000 population). This is visualised in **Figure 1A**.

**Figure 1:**
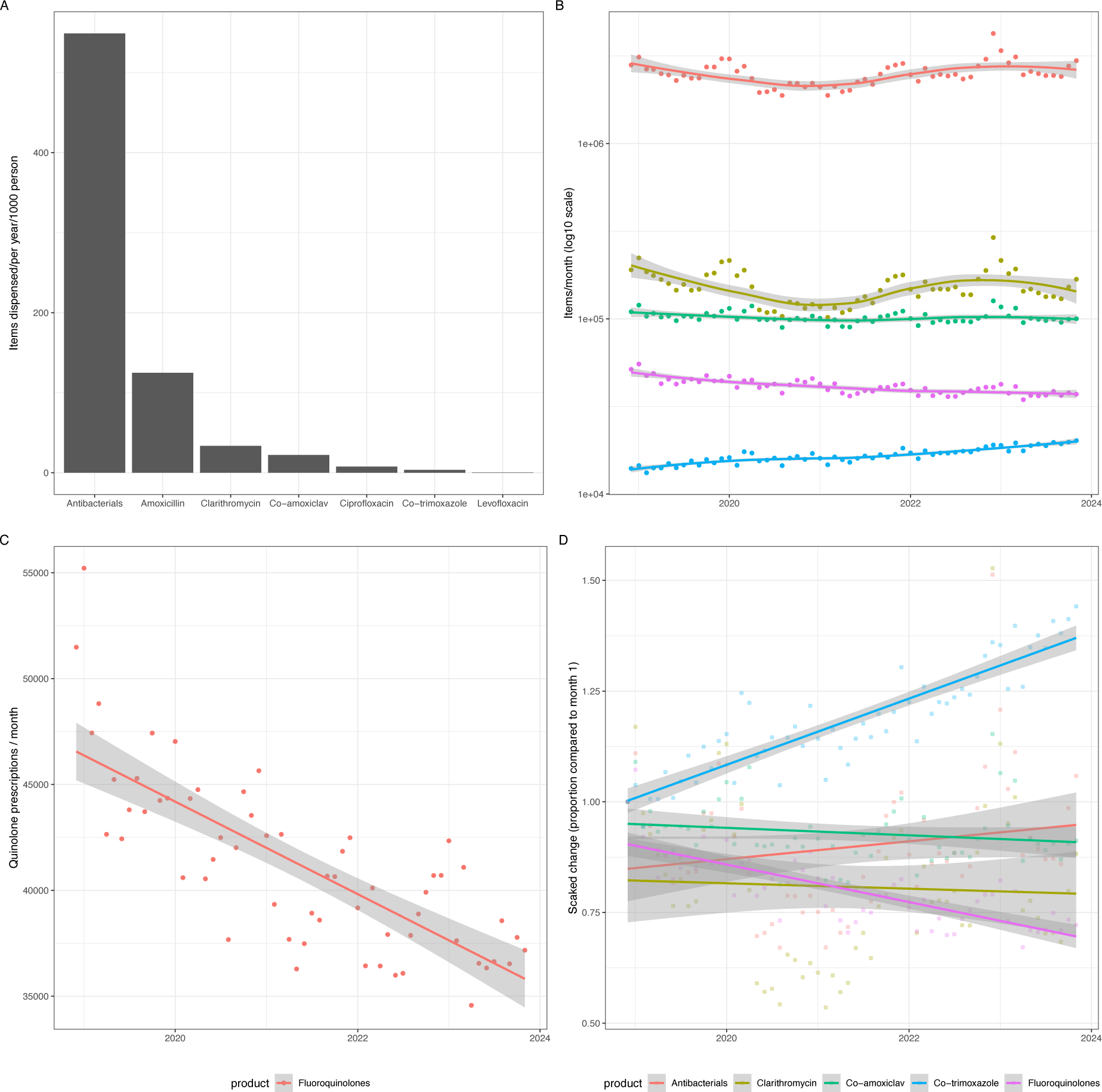
**Panel A** shows the items dispensed per year per 1000 population for a variety of antimicrobials and antimicrobial classes over the whole study period. **Panel B** shows the change over time in prescription of various antimicrobials: clarithromycin, co-amoxiclav, co-trimoxazole, and fluoroquinolones. All antibacterial usage over this time is plotted. **Panel C** shows just fluoroquinolones to help visualise the change, while, **panel D** shows the same as Panel B but rescaled so each drug class starts at 1. Linear regression lines shown with 95% CI.

Over the study period, antibacterial prescription was static (**Figure 1B**, +2% change per year; 95% CI −0.56%; 4.6%, p = 0.12). However, fluoroquinolone prescription decreased substantially over time (−4.2% 95% CI −5.2%; −3.3%, p = 6 x 10^-13^, **Figure 1C**). This was driven by ciprofloxacin reduction, which was 20 times more common than levofloxacin prescription. **Figure 1D** shows these changes scaled so the first month was set to 1 for all products. This change in fluoroquinolone prescribing was offset by a large increase in co-trimoxazole prescribing (change per year +7.5% 95% CI (6.5%; 8.5%), p = 4.4 x 10^-22^). No other tested antimicrobial changed substantially. **Table 1** shows all estimates, with Z-tests comparing change in trends versus all fluoroquinolones. This showed the trend in fluoroquinolone prescription was reduced compared to all other antimicrobials.

**Table 1:**
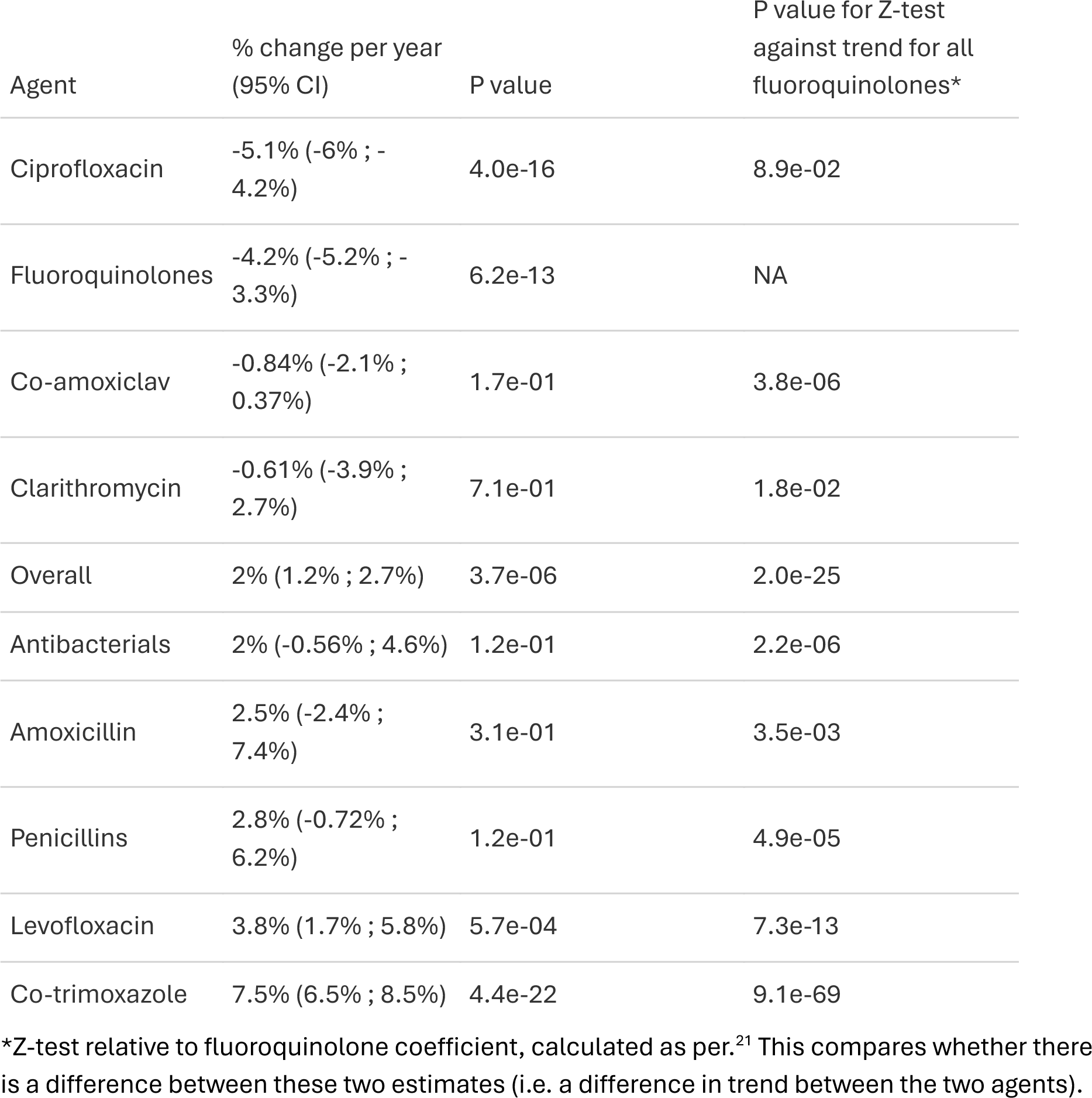
Estimates of the change in prescribing per year of each agent across the primary care study period.

| Agent | % change per year<br>(95% CI) | P value | P value for Z-test<br>against trend for all<br>fluoroquinolones* |
| --- | --- | --- | --- |
| Ciprofloxacin | -5.1% (-6% ; -4.2%) | 4.0e-16 | 8.9e-02 |
| Fluoroquinolones | -4.2% (-5.2% ; -3.3%) | 6.2e-13 | NA |
| Co-amoxiclav | -0.84% (-2.1% ; 0.37%) | 1.7e-01 | 3.8e-06 |
| Clarithromycin | -0.61% (-3.9% ; 2.7%) | 7.1e-01 | 1.8e-02 |
| Overall | 2% (1.2% ; 2.7%) | 3.7e-06 | 2.0e-25 |
| Antibacterials | 2% (-0.56% ; 4.6%) | 1.2e-01 | 2.2e-06 |
| Amoxicillin | 2.5% (-2.4% ; 7.4%) | 3.1e-01 | 3.5e-03 |
| Penicillins | 2.8% (-0.72% ; 6.2%) | 1.2e-01 | 4.9e-05 |
| Levofloxacin | 3.8% (1.7% ; 5.8%) | 5.7e-04 | 7.3e-13 |
| Co-trimoxazole | 7.5% (6.5% ; 8.5%) | 4.4e-22 | 9.1e-69 |

\*Z-test relative to fluoroquinolone coefficient, calculated as per.<sup>21</sup> This compares whether there is a difference between these two estimates (i.e. a difference in trend between the two agents).

There are 101 Integrated Care Boards (ICBs) in England. Over this period, only five care boards reported an increase in fluoroquinolone usage, with only one (West Leicestershire, +3.1% 95% CI, 1.5%; 4.7%, 2.8 x 10^-4^ being measured with any statistical confidence (**Figure 2, Supplementary Table 1**). In contrast, the majority of ICBs had large reductions in fluoroquinolone usage, with 37% of ICBs reducing their prescription by 5% or more per year. The biggest reduction came from NHS Oldham ICB, with a reduction in fluoroquinolone usage by - 12% per year 95% CI (−65%; −47%, p = 1 x 10^-23^).

**Figure 2:**
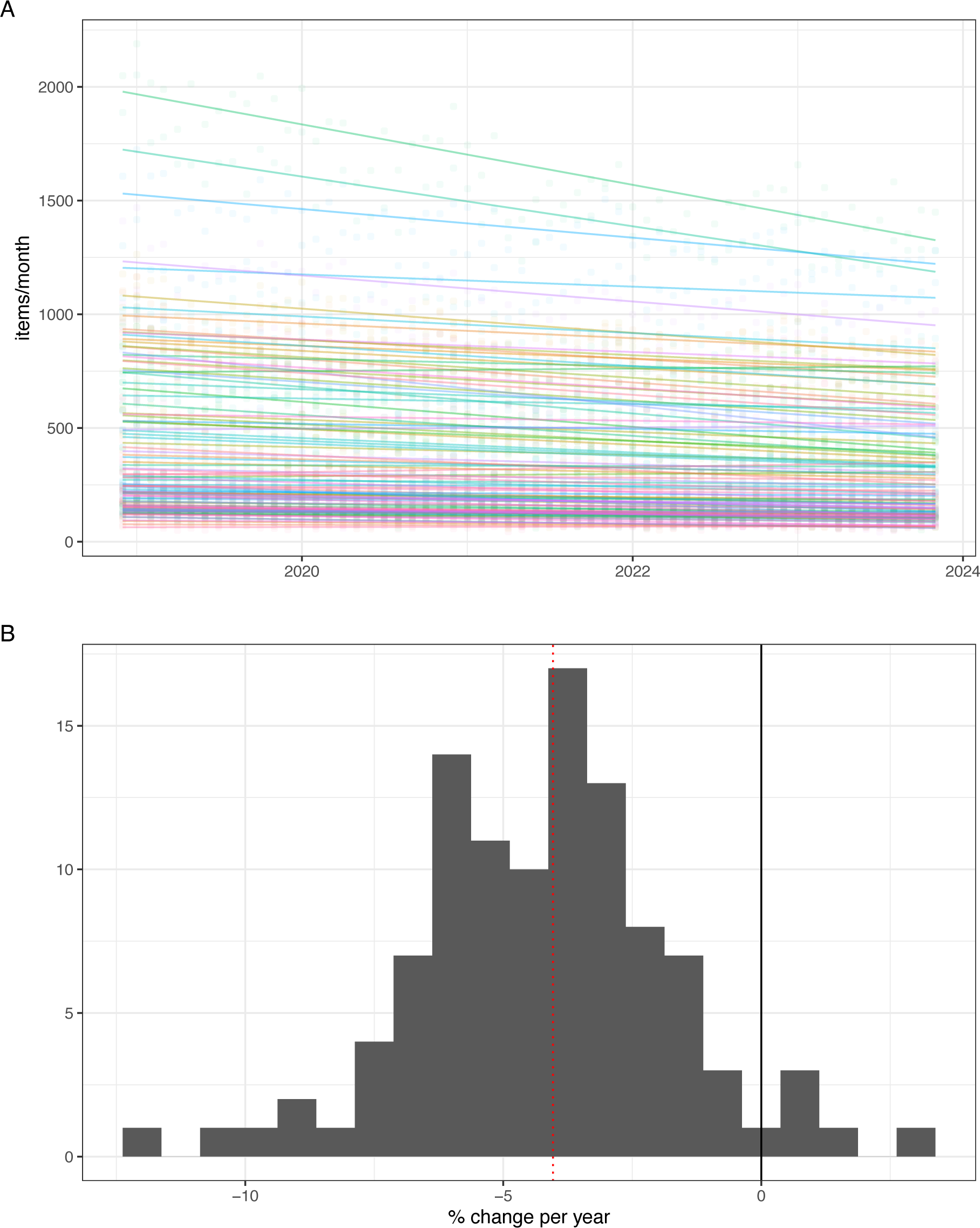
**Panel A** shows changes in prescribing of fluoroquinolones across ICBs. Multiple linear regression lines across all 101 ICBs are fitted. Each line represents an ICB, with the faded dots representing prescribing at that given month. **Panel B** shows the change per year for each ICB plotted as a histogram. The black line represents no change per year, while the dotted red line represents the overall change per year in England.

We then analysed data in secondary care. This is slightly more challenging as the weight of each product in grams is provided, and then this was converted (assumptions in **Methods**) to estimate and compare with the primary care data. In **Figure 3**, we compared estimates for multiple antimicrobial classes in primary and secondary care. Fluoroquinolone usage was generally higher in secondary care (around 50% higher total prescriptions), in contrast to amoxicillin and clarithromycin, which were much more common in primary care. Co-trimoxazole usage was much higher in secondary care. There was much more variability in secondary care prescribing, particularly in early 2020, likely related to variability in hospital occupancy rates the and cancellation of much elective surgery during the COVID-19 pandemic. Unlike in primary care, there were no clear trends identified for ciprofloxacin, clarithromycin, or amoxicillin prescriptions over time in secondary care. However, there was increasing usage of co-amoxiclav, co-trimoxazole, ceftriaxone, levofloxacin, and piperacillin-tazobactam, over this period (all p < 0.001, **Table 2**).

**Figure 3:**
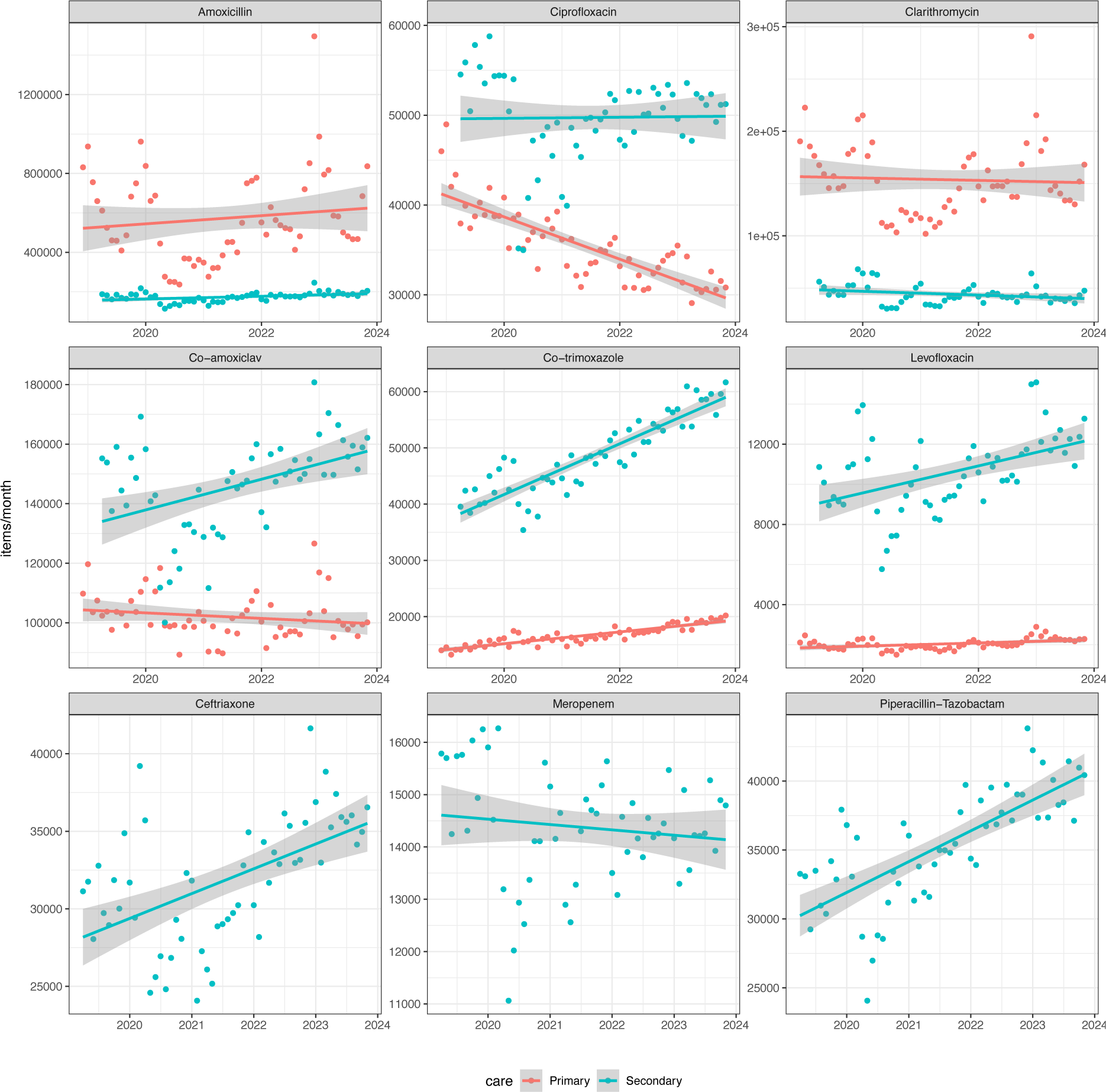
Prescribing (items/month) of various antimicrobial agents in primary and secondary care. Data taken from different sources and transformations applied, so interpret raw comparisons with caution. Linear regression with 95% confidence intervals fitted.

**Table 2:** Relative rates of prescribing of various antimicrobial agents in secondary care. The second column provides the percentage change per year, while the last two columns compare whether the trend is different to the ciprofloxacin or levofloxacin trend.

| Agent | % change per year,<br>(95% CI) | p-value | p-value for Z-test<br>against<br>Ciprofloxacin | p-value for Z-test<br>against<br>Levofloxacin |
| --- | --- | --- | --- | --- |
| Co-trimoxazole | 11% (9.9% ; 13%) | 4.3e-21 | 1.1e-22 | 1.4e-03 |
| Tazobactam | 6.7% (5% ; 8.4%) | 1.3e-10 | 3.3e-08 | 3.9e-01 |
| Levofloxacin | 6.2% (3% ; 9.3%) | 2.3e-04 | 3.6e-04 | 5.0e-01 |
| Ceftriaxone | 5.1% (2.9% ; 7.3%) | 1.9e-05 | 1.8e-04 | 2.9e-01 |
| Amoxicillin | 3.7% (1.4% ; 6%) | 2.5e-03 | 7.1e-03 | 1.0e-01 |
| Co-amoxiclav | 3.3% (1.4% ; 5.2%) | 8.1e-04 | 6.3e-03 | 5.7e-02 |
| Ciprofloxacin | 0.11% (-1.7% ;<br>1.9%) | 9.0e-01 | 5.0e-01 | 3.6e-04 |
| Meropenem | -0.65% (-2% ;<br>0.72%) | 3.5e-01 | 2.5e-01 | 3.2e-05 |
| Clarithromycin | -3.2% (-6.3% ; -<br>0.11%) | 4.3e-02 | 3.1e-02 | 9.8e-06 |

\*Z-test relative to ciprofloxacin coefficient, calculated as per.<sup>21</sup> This compares whether there is a difference between these two estimates (i.e. a difference in trend between the two agents).

Performing a Z-test comparing the regression coefficients, we identified evidence that all these agents had increased prescribing relative to ciprofloxacin prescribing (Z-score p values in **Table 2**). This data suggests that although ciprofloxacin usage has been stable in secondary care (+0.11% 95% CI, −1.7%; 1.9%), the usage of comparable agents with similar spectra of activity has increased relative to ciprofloxacin. Levofloxacin usage did increase over this time period (+6.2% 95% CI, 3%; 9.3%, p = 2 x 10^-4^, but again, relatively less co-trimoxazole and piperacillin-tazobactam (both p <0.001), and with a similar increase to ceftriaxone.

To aid comparison, a final plot showing the ratio of prescribing relative to ciprofloxacin is shown in **Figure 4**. This shows that over the study period, the relative prescribing of ciprofloxacin has fallen. At the start of 2020, approximately 3 prescriptions containing amoxicillin were given for each ciprofloxacin prescription, by late 2023, this was nearly 4. This trend is shown for all tested agents except meropenem, clarithromycin, and levofloxacin, which has a relative increase compared to ciprofloxacin (although small on the absolute terms, of 0.36% per year).

**Figure 4:**
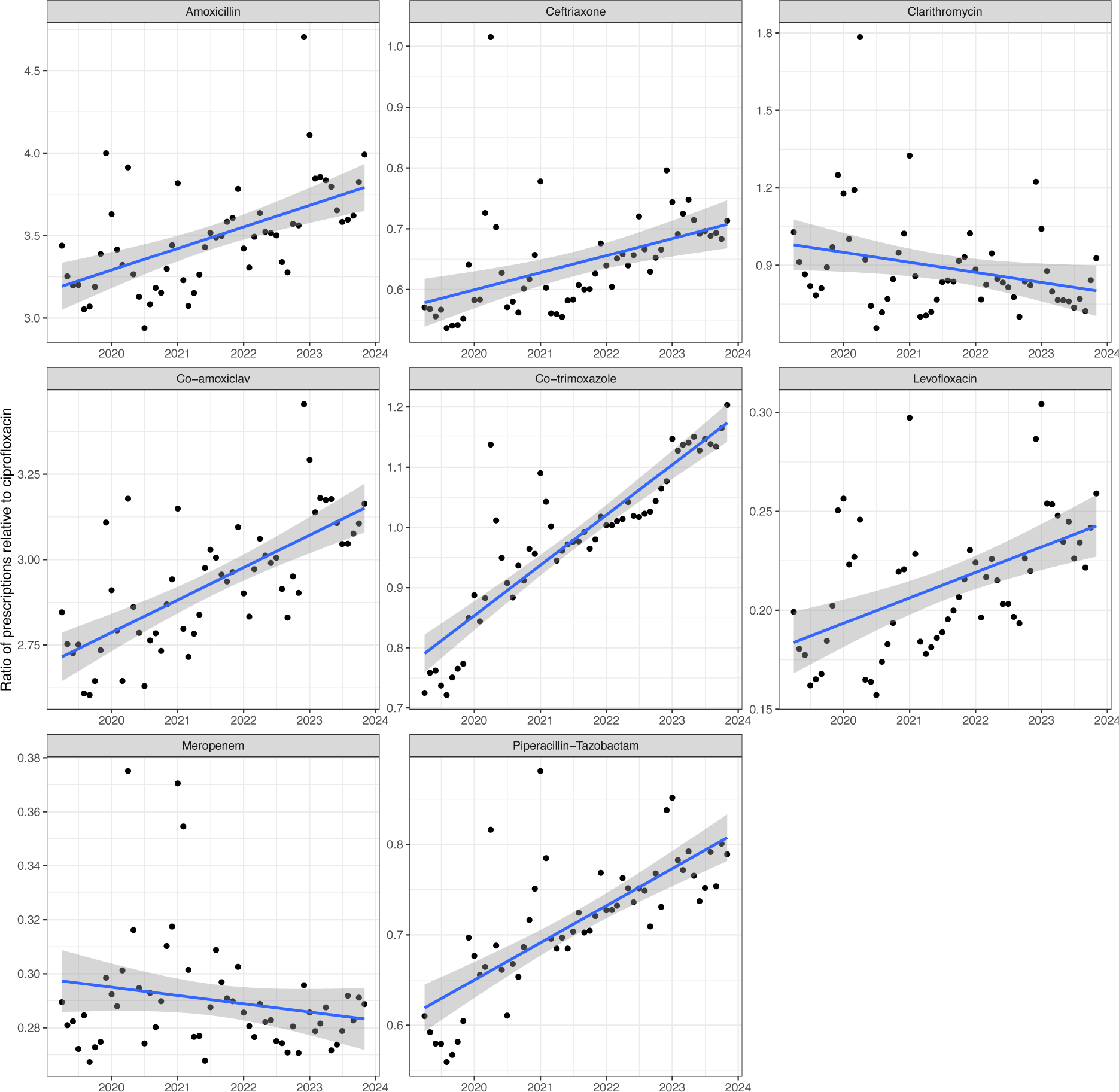
The relative ratio of prescription of each of the above antibacterial agents in secondary care, as compared to ciprofloxacin, over time. Linear regression fitted with 95% confidence intervals.

Finally, from December 2020, we had access to monthly bed occupancy from NHS England and used this to adjust and generate estimated prescriptions per admitted person per month. This period starts >1 year after the MHRA and EMA warnings, and inspection of Figure 3 shows an already steep drop in ciprofloxacin prescribing between 2020 and 2021 of around 15%, which would not be captured in this analysis. Between December 2020 and December 2023, there was a large rise in occupied beds, from around 78,000 to 92,000 inpatients (∼15% increase, **Supplementary Figure 1**). When adjusting for hospital bed utilisation, we identified a reduction in ciprofloxacin (but not levofloxacin usage), with −2% reduction in prescribing per year 95% CI (−3.9%; −0.15%, p = 0.04, **Figure 5**). Given the steep drop prior to this time period this analysis shows that relative to hospital utilisation, reductions in secondary care prescribing are similar, although slight smaller, than in primary care for ciprofloxacin. In contrast, levofloxacin prescription did increase (+4.5% 95% CI +0.63%; +8.3%), meaning overall fluoroquinolone prescriptions were stable (−0.8%; 95% CI −2.5%; +0.8%).

**Figure 5:**
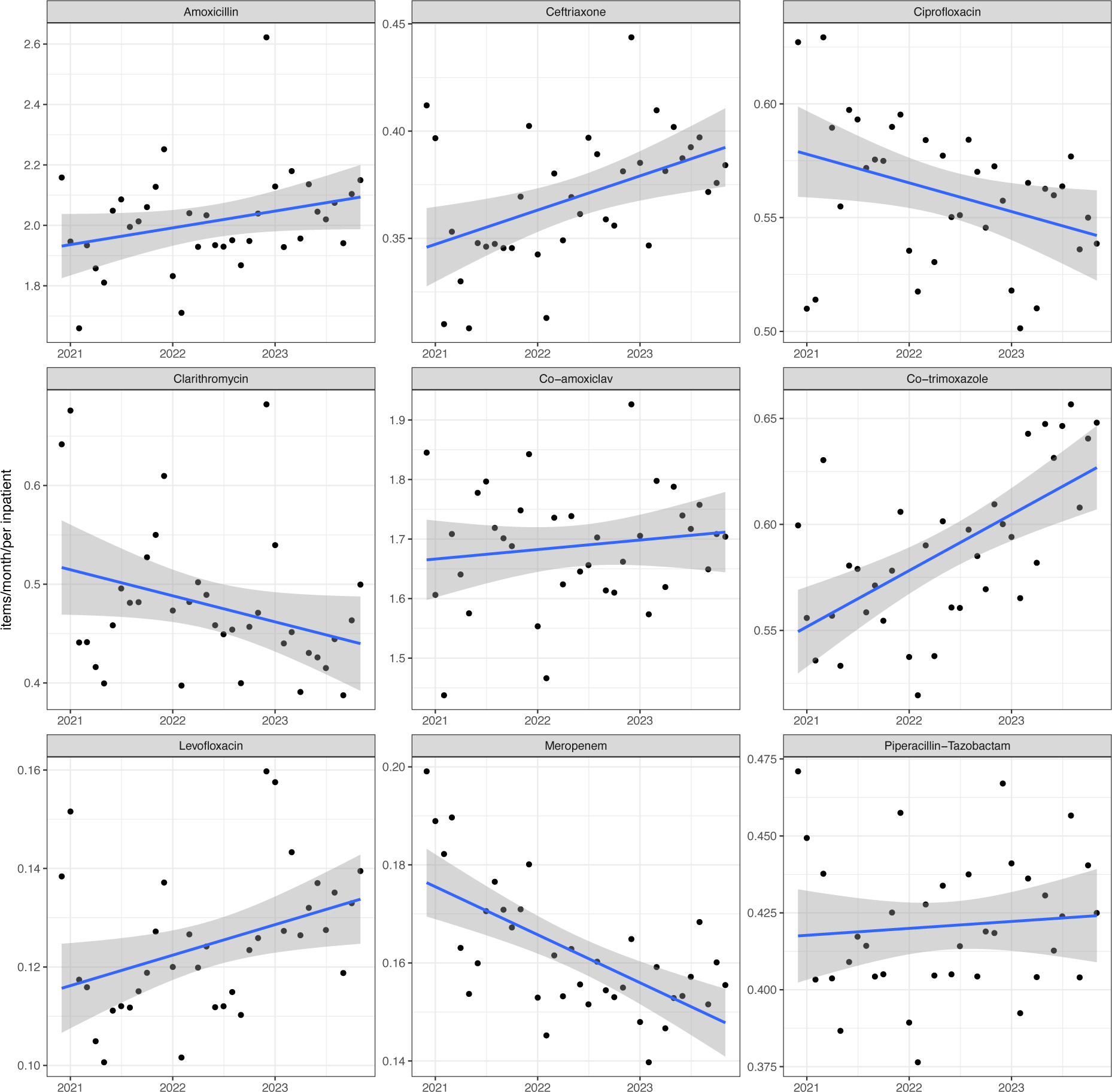
Prescriptions of each agent, adjusted for hospital utilisation, so the Y-axis is on the scale of prescriptions per inpatient per month. Linear regression (blue line) fitted with 95% CI (grey area)

## Discussion

Our work uses both primary and secondary care data for the whole of England to explore fluoroquinolone prescriptions over the last five years and assess the influence of EMA/MHRA restrictions on prescribing. We identified clear reductions in primary care prescribing of ciprofloxacin, which accounted for the vast majority of all primary care fluoroquinolone prescriptions. This was in contrast to usage of comparator antibiotics that were largely static.

In secondary care, antimicrobial prescribing varied much more, likely due to the COVID-19 pandemic. Absolute ciprofloxacin prescribing was stable across the time period. However, comparator antibiotic prescriptions increased, consistent with a relative reduction in ciprofloxacin prescribing. For the time period from December 2020 onwards (by which time a ∼15% reduction in ciprofloxacin prescribing had occurred), we had hospital utilisation data. When adjusting for this, relative ciprofloxacin usage reduced by around 2% per year. This is consistent with increased pressure and workload on the NHS in England, while maintaining relatively infrequent use of fluoroquinolones compared to other agents ?. Levofloxacin prescription did, however, increase.

One major reason for the continued use of fluoroquinolones in secondary care is that there are remarkably few good oral options for the therapy of some infections. Potential agents often require intravenous administration, and there is also pressure to reduce the use of certain other agents (e.g. third generation cephalosporins, Beta-lactam-beta-lactamase inhibitors and carbapenems). Resistance rates to co-trimoxazole and co-amoxiclav in Gram negatives are ∼30%, with ciprofloxacin at only 17% in the recent ESPAUR report.^24^ Additionally, some randomised trial data and meta-analyses, particularly in pneumonia, suggests that fluoroquinolones are superior to other options in this condition, which is the main reason for antimicrobial prescribing in acute admissions.^25^ Other indications for fluoroquinolones may be relatively uncommon in terms of proportional numbers of patients affected, but by their nature require prolonged courses for up to 3-6 months, for example prosthetic joint or vascular implant infection.

As far as we are aware, the only other study examining post EMA/MHRA fluoroquinolone prescribing trends is Ly et al, funded by the EMA and analysing UK data from 2016-2020^15^. This study used a multisegmented regression approach to identify monthly percentage changes in relation to specific communications from regulators. Although this study is reported in the MHRA Drug Safety Update as showing ‘no change’ in prescribing, it does show a reduction in fluoroquinolone prescribing, and is estimated by Ly et al to be “at best, a reduction in prescriptions of around 25%”.^15^ As such, our results are concordant and extend the study period and show further reductions since then. The English Surveillance Programme for Antimicrobial Utilisation and Resistance (ESPAUR) report for 2023 provides supportive data, with quinolone prescription dropping from 0.565 in 2018 to 0.456 in 2022 (p = 0.04).^24^ Interestingly, a study of all antimicrobial prescribing over a longer period (2014-2024), identified a reduction in overall antibiotic prescribing, whereas our data suggested largely static antimicrobial prescribing.^26^

Limitations of our study include its entirely ecological focus. We had no access to individual level data, and so we can only focus on trends without deeper insight. Interpretation of our secondary care data was reliant on transformation of product weight into assumed product courses, although these transformations would only change absolute differences between comparator antibiotics, and not changes in trend. Our use of hospitalisation data as a proxy for hospital pressure is imperfect, and only reflects total inpatients (not, e.g. admissions). Given fluoroquinolones are used in both acute admissions and inpatients, it would be challenging to accurately model hospital pressure, and inpatient hospitalisation is a reasonable proxy.

In summary, national prescribing data for England shows an approximate 5% reduction per year in ciprofloxacin prescribing in primary care, where there is the most concern about ‘inappropriate’ prescribing of this drug class. This is clearly a large, statistically precise, and important reduction, which has continued over the 5-year period however the exact reasons for this reduction cannot be determined from this data.

## Data Availability

We used R v 4.31 for analyses, and used the tidyverse package for data wrangling and plotting.22 All code is available at https://github.com/gushamilton/floroquinolone, and the analysis is completely replicable by running the same code locally. Regression was performed using the linear regression model in R, while some plots fitted splines via ggplot.

## Supplementary Figures

**Figure S1:**
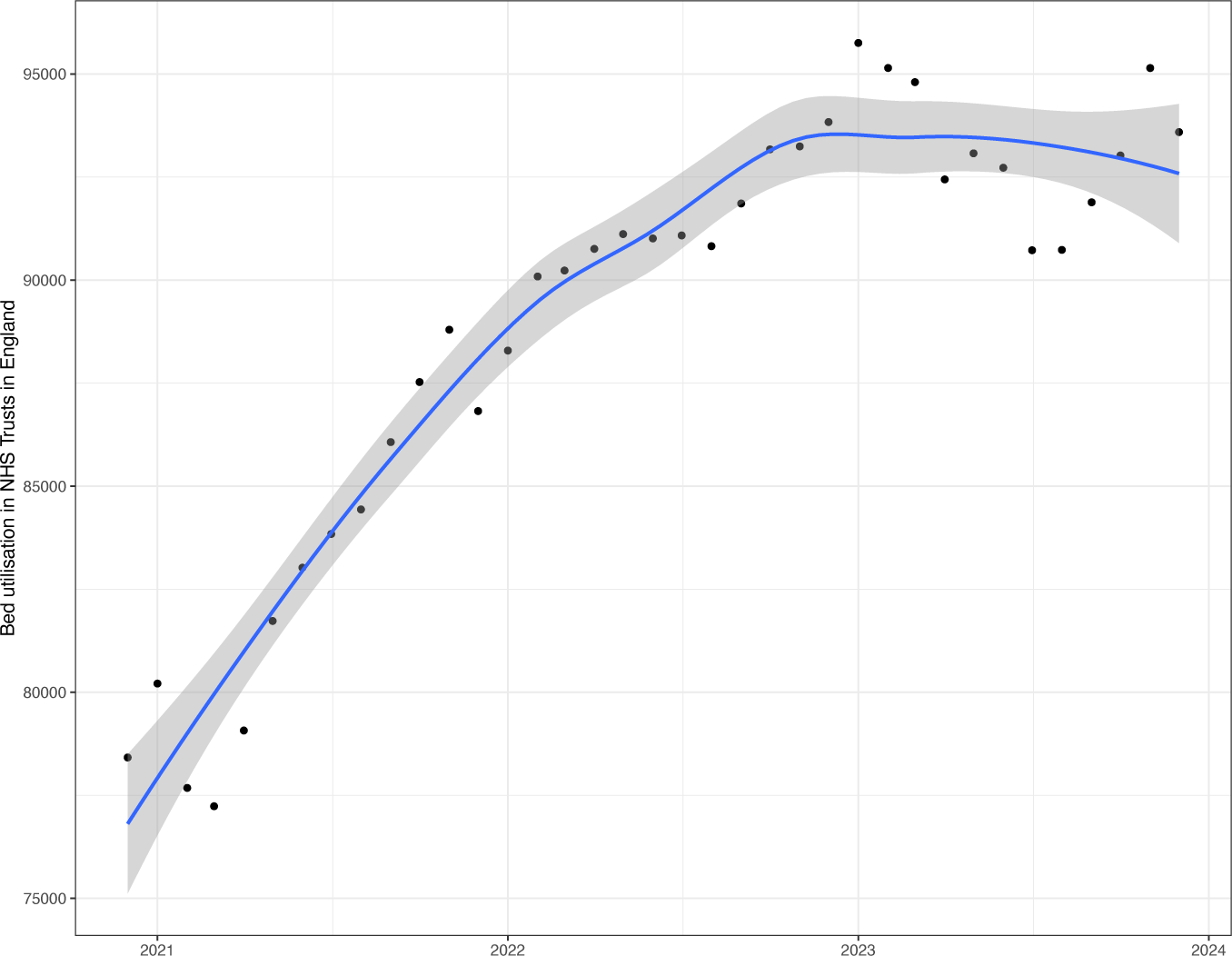
Bed utilisation in English NHS trusts from December 2020 until December 2023. Data from NHS England.

